# Tractography-guided versus Clinical Contact Selection for Deep Brain Stimulation in Tremor - A Prospective Clinical Trial

**DOI:** 10.1101/2025.05.30.25328382

**Authors:** Christina van der Linden, Thea Berger, Till A. Dembek, Gregor A. Brandt, Joshua N. Strelow, Juan Carlos Baldermann, Hannah Jergas, Gereon R. Fink, Veerle Visser-Vandewalle, Michael T. Barbe, Jan Niklas Petry-Schmelzer

## Abstract

**Background:** Imaging-guided programming strategies for deep brain stimulation (DBS) offer the potential to reduce programming time and complexity. However, evidence-based paradigms tailored to tremor remain unavailable. Converging research points to an involvement of the dentato-rubro-thalamic tract (DRTT) in tremor pathophysiology across diseases, providing a potential target for imaging-guided programming in Essential Tremor (ET) and Parkinson’s Disease (PD).

**Objective:** To prospectively evaluate whether imaging-guided contact selection for tremor control based on individual tractography of the DRTT is non-inferior to clinical contact selection.

**Methods:** Tremor control was assessed in an acute challenge for each DBS contact in 16 ET and 24 PD patients with directional DBS leads in the thalamic ventral intermediate nucleus/ posterior subthalamic area (ET) or subthalamic nucleus (PD). The effect of the clinically most effective contact was compared to the effect of the contact with the largest overlap of the stimulation spread with the individual DRTT. Primary outcome was the difference in relative tremor improvement between clinical and imaging-guided contact selection, assessed by an accelerometry-based tremor score, assuming a non-inferiority margin of ≤ 20%.

**Results:** Tremor control achieved via the imaging-based selected contact was non-inferior to tremor control via the contact chosen by clinical testing for ET (median difference best clinical vs. best imaging-based contact-8%, 95% CI-11.5% to-3.5%) and PD (median difference - 10%, 95% CI-20% to-4%).

**Conclusion:** This study provides class-II evidence that the overlap of the stimulation spread with the individual DRTT is a suitable marker for imaging-guided contact selection for DBS in tremor.

## Introduction

Tremor is one of the most prevalent and disabling symptoms in movement disorders, primarily caused by Parkinson’s Disease (PD) and Essential Tremor (ET). Deep brain stimulation (DBS) is a well-established treatment for medication-refractory tremor, targeting the subthalamic nucleus (STN) for PD and the thalamic ventral intermediate nucleus (VIM) and posterior subthalamic area (PSA) for ET [1,2]. Postoperative tremor control crucially depends on precise contact selection. Especially with novel DBS devices offering vertical and horizontal current steering with at least eight contacts per lead, the complexity of trial-and-error-based clinical contact selection dramatically increased. Therefore, time-and cost-efficient programming approaches such as imaging-guided strategies gained increasing importance [3–5].

A potential target for imaging-guided contact selection for tremor is the dentato-rubro-thalamic tract (DRTT), a central pathway of cerebello-thalamo-cortical networks, which has been suggested to be involved in tremor pathophysiology across diseases [6,7]. The DRTT originates from the cerebellar dentate nucleus and ascends via the superior cerebellar peduncle, with approximately 90% of the fibers crossing to the contralateral side at the level of the pons. It further passes through the red nucleus, medially past the STN, and then terminates within the thalamic nucleus ventralis oralis posterior and the VIM, from where neurons mainly project to the primary motor cortex [8]. While the inferior border of the VIM has been the target of choice for neuromodulation in ET for years, recently, attention has shifted to the region inferior and posterior to the VIM, the PSA. The superior efficiency of PSA stimulation over VIM stimulation was shown in two randomized, cross-over studies [1,9], and subsequent studies confirmed that these findings were related to the closer proximity of PSA contacts to the DRTT [10]. Other studies investigated direct surgical targeting of the DRTT for DBS in ET [11], and also across tremor diseases [12]. In a recent retrospective pilot study, we explored an imaging-guided contact selection approach for tremor control based on the overlap of the stimulation spread with the DRTT in a small group of patients with ET [13]. Regarding Parkinsonian tremor, previous work indicates that the tremor-suppressive effect of STN-DBS might also be associated with stimulation of the DRTT, with active contacts closer to the DRTT being more effective for tremor suppression [14,15], underlining the crucial role of cerebello-thalamo-cortical networks in the “dimmer-switch-model” of Parkinsonian tremor [16].

Taken together, these findings led to the hypothesis that modulation of cerebello-thalamic networks through DRTT stimulation is associated with tremor control across diseases and DBS targets [7,17,18]. Consequently, this clinical trial aimed to prospectively evaluate the hypothesis that contact selection for tremor control based on standardized individual tractography of the DRTT is non-inferior to clinical contact selection in patients with PD and ET in an acute challenge.

## Patients and Methods

### Patient Selection and Ethics

The *TremTract Study* was a prospective single-center double-blinded clinical trial. Patients aged ≥ 18 and ≤ 80 years with a confirmed diagnosis of ET or PD according to the International Parkinson and Movement Disorder Society consensus diagnostic criteria were eligible for study participation [19,20]. Patients underwent uni-or bilateral implantation of directional DBS leads in VIM/PSA (ET) or STN (PD) at least three months before study participation. Only patients with a relevant tremor burden in medication OFF and stimulation OFF condition in at least one upper extremity were included. For ET, a relevant tremor burden was defined as ≥ 2 of 4 points in one of the items 5 and 6 of the Fahn Tolosa Marin Tremor Rating Scale (FTM-TRS, postural or action/intention tremor) [21]. For PD, a relevant tremor burden was defined as ≥ 2 of 4 points in item III.17 of the MDS-Unified Parkinson’s Disease rating scale (MDS-UPDRS III, rest tremor amplitude) [22]. Only leads implanted in the hemisphere contralateral to the body side with relevant tremor burden were tested. Additional inclusion criteria comprised the availability of imaging data for lead reconstruction and fiber tracking, i.e., preoperative anatomical T1 and T2-weighted MRI sequences, diffusion tensor imaging, and postoperative CT imaging. All patients underwent DBS surgery at our center as per clinical routine after a multidisciplinary DBS board evaluation, which was unrelated to this study. The local ethics committee of the University of Cologne approved the study (21-1441), conducted under the Declaration of Helsinki, and registered with the German Clinical Trials Registry before patient recruitment (DRKS00026596). All patients gave written informed consent before participation. Clinical testing was conducted at the Department of Neurology of the University Hospital Cologne between 02/2022 and 01/2023.

### Determination of the Best Clinical Contact

Figure 1 summarizes the study design. Clinical assessments were performed after withdrawal of dopaminergic or tremor-suppressing medication at least 12 hours before examination (MedOFF). First, the overall stimulation effect on motor symptoms was determined by assessing the UPDRS-III for patients with PD and the FTM-TRS for patients with ET in MedOFF/Stimulation ON (StimON) and MedOFF/Stimulation OFF (StimOFF) state after a stimulation wash-out period of 20 minutes. Then, a contact-wise monopolar review was conducted with the stimulation frequency set to 130 Hz, the pulse width set to 60 µs, and the contralateral stimulation switched off. Contacts and, if applicable, hemispheres were tested in randomized order, and in the case of directional levels, each directional segment was investigated separately. Clinical testing was conducted (i) once per lead at baseline (StimOFF), (ii) at a fixed predefined amplitude (VIM/PSA-DBS: 1.5 mA, STN-DBS: 2.0 mA), and (iii) at the maximally tolerated amplitude, i.e., 0.5 mA below the threshold of stimulation-induced side effects. The occurrence of side effects was evaluated with amplitude increments of 0.5 mA. Clinical testing was conducted only once at the maximal tolerated amplitude if a contact’s side-effect threshold was below the fixed predefined amplitude (see Supplementary Figure S1 for an overview of the clinical testing algorithm). A fixed amplitude of 2 mA was chosen for PD based on clinical experience and is commonly used in monopolar reviews [23]. As observed previously, in ET, a stimulation with 2 mA often led to complete tremor suppression by multiple contacts [13]. Hence, to improve the discrimination of distinct differences in tremor effects per contact, a reduced fixed amplitude of 1.5 mA was chosen for the ET cohort.

**Figure 1:**
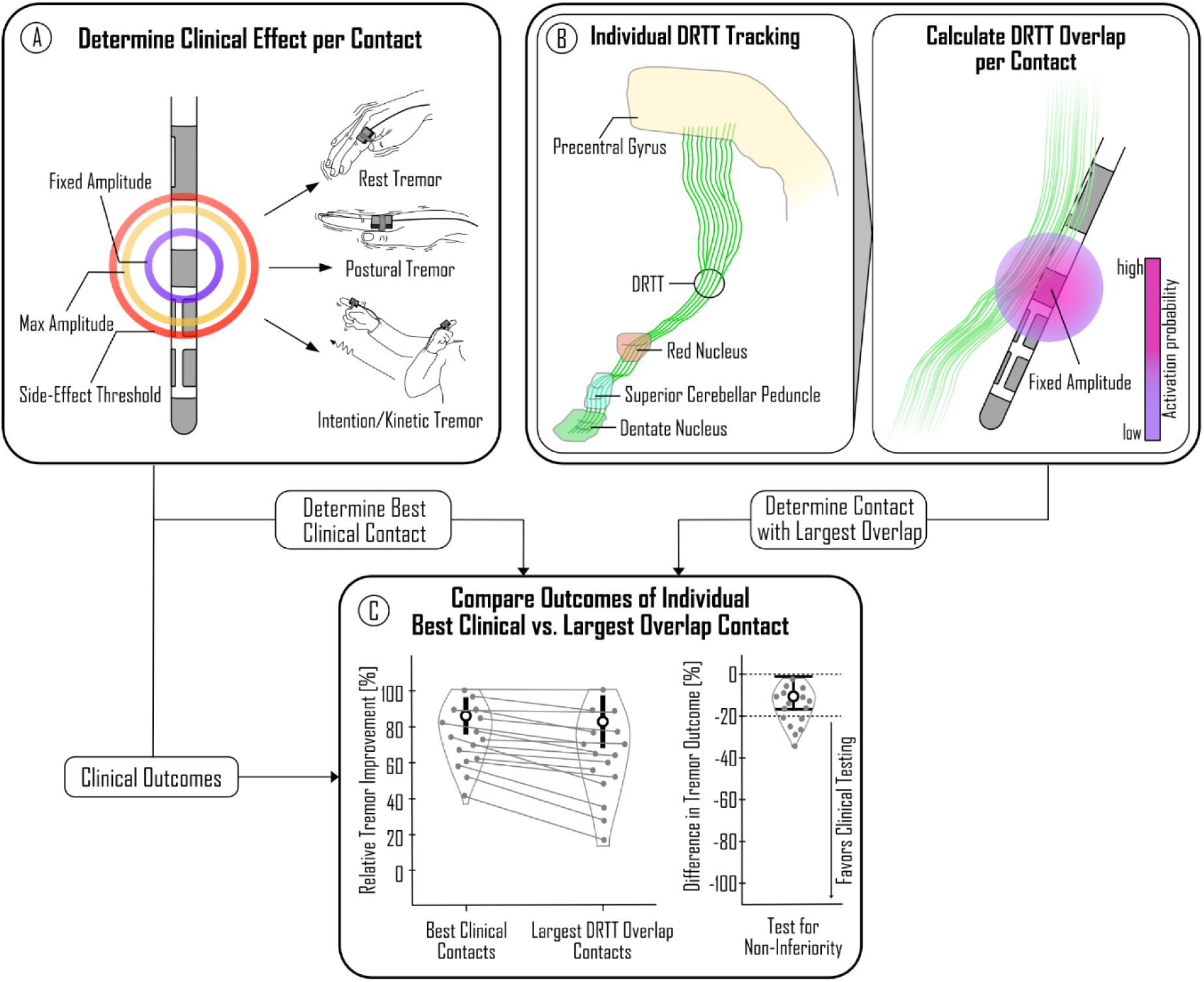
Study Design. (A) The clinically most effective stimulation contact was determined in all patients by clinical testing of rest, postural, and intention/kinetic tremor in a randomized order and double-blinded experimental setup. (B) The best imaging-guided contact was selected based on the stimulation overlap with the individual DRTT. (C) As primary outcome, the difference in relative tremor improvement, achieved by the best clinically chosen and imaging-guided contacts with the predefined fixed amplitude, was tested for non-inferiority with a non-inferiority margin of 20%. Abbreviations: DRTT = dentato-rubro-thalamic tract

Clinical testing included the assessment of rest, postural, and kinetic tremor following the instructions of MDS-UPDRS III items III.15, III.16. and III.17 with standardized video-based instructions (PsychToolbox Version 3.0.19 for MATLAB) [24,25]. For intention/kinetic tremor, patients were asked to press a button, reachable with the arm outstretched in front of them, and move the arm back to wing position alternatingly. Each task was performed for 20 seconds, with each trial’s beginning and end indicated by an acoustic signal. Tremor severity was measured by triaxial accelerometry at the index finger and wrist (Supplementary Material 1.1). Additionally, tremor severity was rated by an experienced clinician (CvdL) blinded to the current stimulation settings based on the tremor amplitude cutoffs provided by MDS-UPDRS III Items III.15–III.17. In patients with PD, akinetic-rigid symptoms were evaluated based on MDS-UPDRS III items III.3 (Upper Extremity Rigidity) and III.4 (Finger Tapping).

A stepwise selection process defined the lead-wise best clinical contact. Contacts were first ranked by the highest percentage improvement of the accelerometric tremor score at the fixed amplitude. If multiple contacts showed equal effectiveness at the fixed amplitude, they were further sorted by the highest percentage improvement at the maximal amplitude and then by the highest side effect threshold.

### Determination of the Best Imaging Contact

#### Probabilistic Tracking of the DRTT

All patients underwent routine MR-imaging for surgical planning. Diffusion data processing was performed using the MRtrix3Tissue package (https://3Tissue.github.io), a fork of MRtrix3 [26]. Probabilistic fiber tracking of the DRTT was conducted with the contralateral dentate nucleus as seed region, the contralateral superior cerebellar peduncle, the ipsilateral red nucleus, and the ipsilateral precentral gyrus as waypoints (Figure 1B) and 500 selected fibers [10–13,27]. Regions of interest were delineated in the MNI ICBM 2009b asymmetric template to ensure a standardized tracking result with reduced inter-rater variability. Afterwards, they were transformed to the preoperative T1 space using Advanced Normalization Tools (ANTs), and further co-registered to the averaged b0 using SPM (http://www.fil.ion.ucl.ac.uk/spm/software/spm12/), as described previously [10,13]. Finally, the resulting tracts were co-registered to the preoperative T1. More details on the diffusion imaging analysis are provided in Supplementary Material 1.2.

#### Calculation of Contact-Wise DRTT Overlap

DBS leads were reconstructed from postoperative CT scans and transformed into T1 space utilizing a well-established workflow in the LEAD-DBS toolbox (version 3, www.lead-dbs.org, Supplementary Material 1.3) [28]. The local stimulation spread at the fixed amplitude was calculated in T1 space for each contact. To do so, precomputed electric fields were employed for each stimulation setting to estimate the spread of the electric field for homogenous tissue with a conductivity of σ = 0.2 S/m (FastField) [29,30]. Instead of applying a fixed electric field threshold resulting in a binarized stimulation volume, a recently established probabilistic stimulation model based on a sigmoidal activation function and established electric field activation thresholds for different fiber diameters was used, resulting in a voxel-wise activation probability value reaching from 0 (= no activation) to asymptotic 1 [31]. Ultimately, the overlap of each stimulation spread with the respective DRTT was calculated as the sum of the fiber-wise maximal activation probability of each fiber, stimulated with an activation probability of at least 0.05. Contacts were ranked lead-wise by overlap, declaring the contact with the highest overlap as the best imaging contact. Investigators performing the tracking and overlap analysis (JNPS) were blinded to the clinical testing results and vice versa.

### Primary and Secondary Outcome Measures

The primary outcome measures were the differences in relative improvement of the accelerometric total tremor score at the fixed amplitude by the best clinical contact and the best imaging-guided contact for ET and PD respectively. The accelerometric tremor score was derived by modeling the non-linear relationship between the mean acceleration captured by accelerometry and the clinical rating, allowing for an objective, continuous tremor rating (Supplementary Material 1.1), which was extensively validated in a prior study [32]. The accelerometric total tremor score was calculated as the sum of all accelerometric tremor subscores (rest, postural, and intention/kinetic tremor) clinically rated with more than 0 point at the respective baseline.

Secondary outcomes were (i) differences in relative improvement of the accelerometric tremor subscores (ET: postural and intention tremor, PD: rest tremor and action tremor, calculated as the sum of postural and kinetic tremor) at the fixed amplitude, (ii) differences in relative tremor improvement at the maximum tolerated amplitude, (iii) difference in side-effect thresholds, and (iv) duration of clinical assessment vs. processing of imaging data respectively. Furthermore, for patients with PD, the difference in the relative improvement of akinetic-rigid symptoms was evaluated, based on the sum score of the MDS-UPDRS III items III.3 (Upper Extremity Rigidity) and III.4 (Finger Tapping) [22]. Ultimately, we conducted a pooled analysis to compare the difference in the primary outcome across diseases and target points.

### Sample Size Calculation

A power analysis was conducted to test the primary outcome in a non-inferiority analysis with a non-inferiority margin of 20% [33,34], a one-sided type I error of 2.5%, and a power of 90% for ET and PD separately. The required sample size for ET was calculated based on a pilot study by our group, which resulted in a sample size of 28 hemispheres [13]. Since there was no pilot study available for PD, sample size calculation for PD relied on a recent study by Waldthaler et al. comparing imaging-guided and clinical programming strategies for akinetic-rigid symptoms in STN-DBS, resulting in a total of 28 hemispheres as well [33].

### Statistical analysis

For non-inferiority analyses, we calculated the 95% confidence interval (CI) of the median difference between the best clinical and the best imaging contact via bootstrapping with n = 10000 iterations. Non-inferiority was assumed if the lower margin of the 95% CI was ≥-20%. For the secondary outcomes, the Wilcoxon signed-rank test was employed for group comparisons. To investigate the relationship between relative tremor improvement and the overlap with the DRTT, linear mixed effect models were calculated, implementing lead as the random effect to correct for multiple testing per lead (Tremor improvement ∼ 1 + Overlap + (1|lead)). Data distribution was assessed by the Shapiro-Wilk-Test. Where applicable, p-values are reported to a significance level of 0.05. All analyses were conducted in MATLAB Version R2023a (The MathWorks Inc., Natick, Massachusetts, United States).

## Results

### Patient Characteristics

We included 16 patients with ET (N = 28 hemispheres) and 24 patients with PD (N = 29 hemispheres) (Figure 2). Clinical testing was completed for all but one hemisphere. Supplementary Table 2 summarizes the patient characteristics, and lead locations are illustrated in Figure 3A (ET) and 4A (PD).

**Figure 2:**
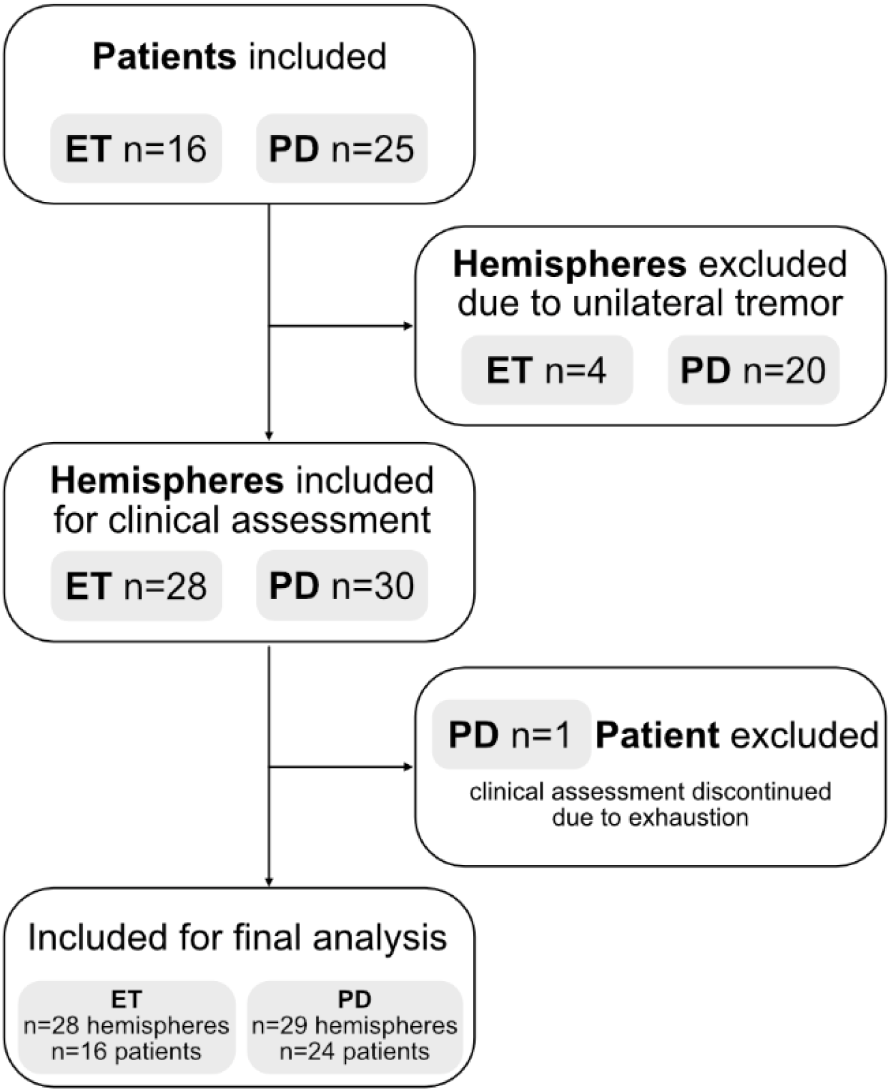
Patient Selection. 16 patients with ET were included, all with bilateral VIM/PSA leads, but only 12 with bilateral tremor in StimOFF, resulting in 28 included hemispheres. For PD, 24 patients were included, 20 with unilateral tremor, and 5 with bilateral tremor, resulting in 30 hemispheres included for clinical assessment. 1 patient with unilateral tremor was excluded because auf exhaustion during the clinical assessment. Abbreviations: ET = Essential Tremor, PD = Parkinson’s Disease

**Figure 3:**
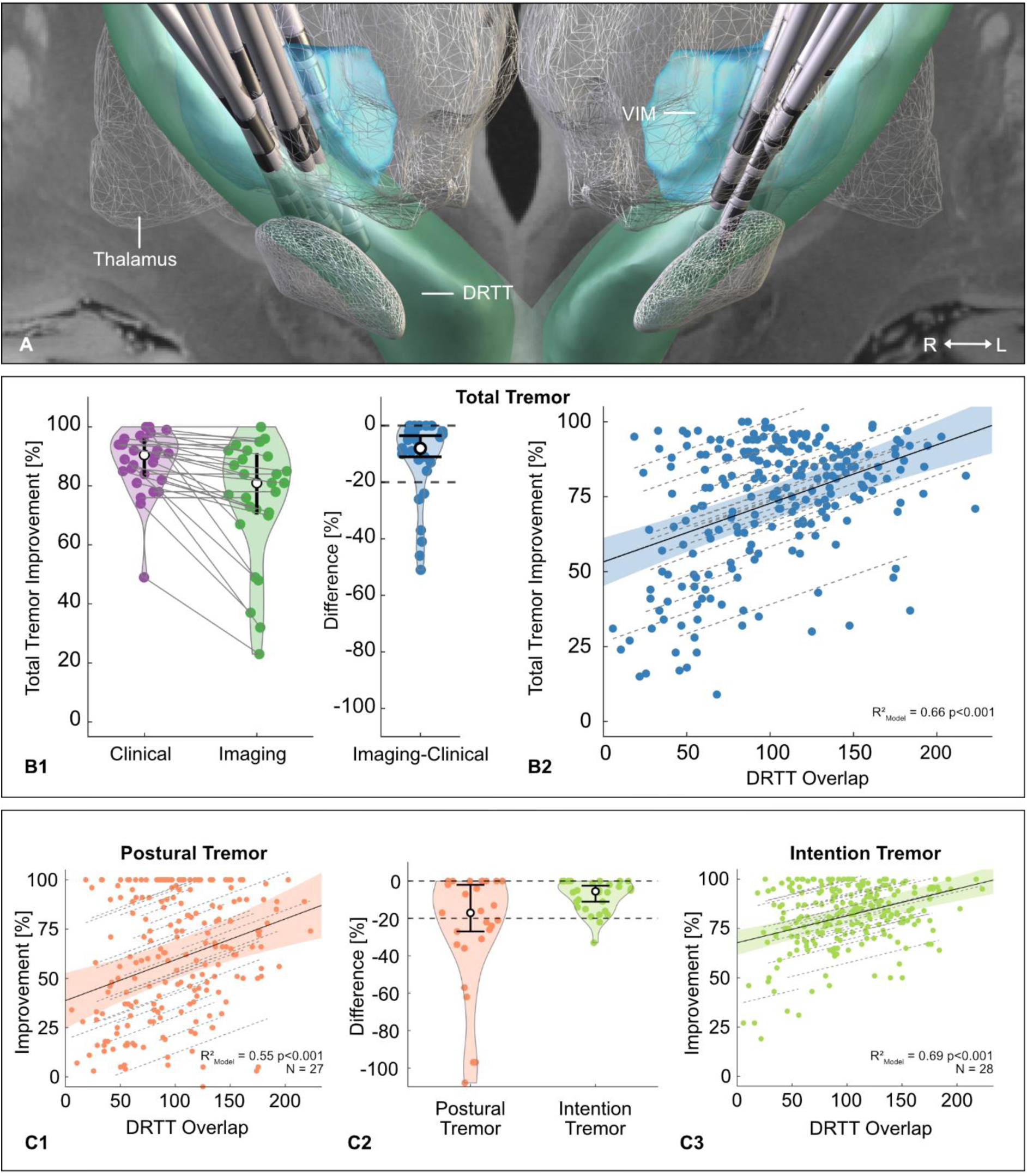
Results of the Essential Tremor Cohort. (A) Lead positions in MNI ICBM 2009b space, illustrated with relevant anatomical structures and a study cohort-based common DRTT. (B1) Total tremor control by stimulation via the imaging-guided selected contact was non-inferior to the best clinical contact. (B2) A strong association between total tremor improvement and the DRTT overlap was observed. (C) Tremor Subscores. Non-inferiority was established for intention tremor but not postural tremor (C2). Nevertheless, a strong association existed between the respective improvement of tremor subtypes and the DRTT overlap (C1&C3). Abbreviations: DRTT = dentato-rubro-thalamic tract, PSA = posterior subthalamic area, STN = subthalamic nucleus, VIM = ventral intermediate nucleus

### Primary Outcomes - Difference in Total Tremor Control

In ET, the median improvement of the total tremor score achieved by the best clinical contact was 90.5% (IQR 83% to 96%) and 81% (IQR 71% to 91%) by the best imaging-guided contact. The median difference in tremor control between the contact selection strategies was-8% (IQR-14% to-2%) with a 95% CI of-11.5% to-3.5%, indicating non-inferiority (Figure 3B, Table 1). In PD, the median improvement achieved by stimulation via the best clinical contact was 81% (IQR 65% to 93%) and 64% (IQR 31% to 83%) for the best imaging-guided contact. The median difference in tremor control between the contact selection strategies was-10% (IQR-22% to-0.02%) with a 95% CI of-20% to-4% (Figure 4B, Table 1), also indicating non-inferiority.

**Figure 4:**
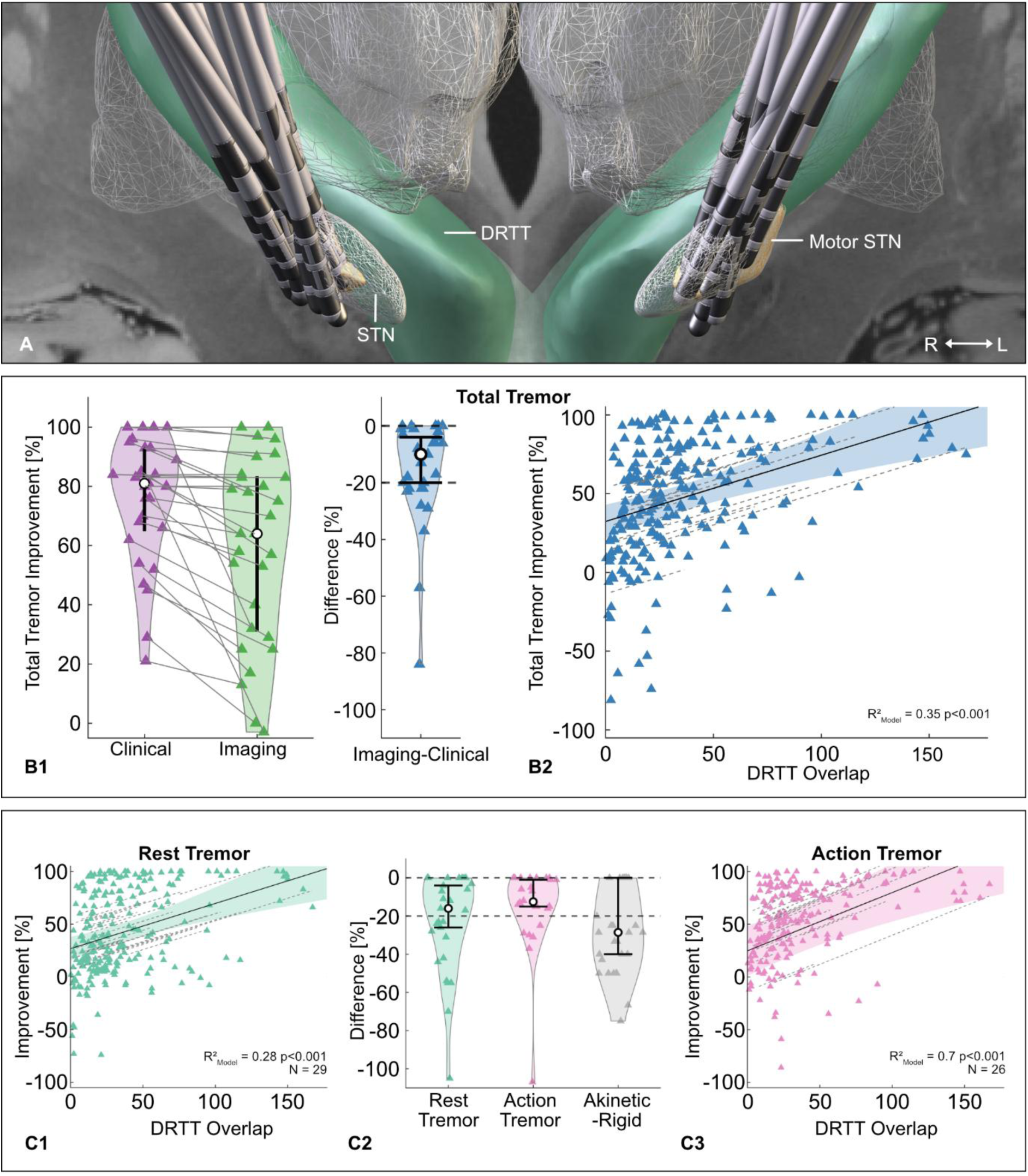
Results of the Parkinson’s Disease Cohort. (A) Lead positions in MNI ICBM 2009b space, illustrated with relevant anatomical structures and a study cohort-based DRTT. (B1) Total tremor control by stimulation via the imaging-guided selected contact was non-inferior to the best clinical contact. (B2) A moderate association between total tremor improvement and the DRTT overlap was observed. (C) Tremor subscores. Non-inferiority was achieved for action tremor but not rest tremor or akinetic-rigid symptoms (C2). The association between DRTT overlap and tremor improvement was stronger for action tremor than rest tremor (C1&C3). Abbreviations: DRTT = dentato-rubro-thalamic tract, PSA = posterior subthalamic area, STN = subthalamic nucleus, VIM = ventral intermediate nucleus

**Table 1.** Tremor Improvement by Clinical and Imaging-based Contact Selection at the Fixed Amplitude. Relative improvement of the accelerometric tremor score achieved by the best clinical contact and the best imaging-guided contact with the fixed amplitude (1.5 mA for ET, 2.0 mA for PD) and median of the differences between the improvements. Abbreviations: CI = Confidence Interval, ET = Essential Tremor, IQR = interquartile range, PD = Parkinson’s Disease

| Outcome parameter | N | Median Improvement Clinical [IQR] | Median Improvement Imaging [IQR] | Median Difference Clinical – Imaging [IQR] | 95% CI |
| --- | --- | --- | --- | --- | --- |
| **ET** |  |  |  |  |  |
| **Total Tremor** | **28** | **0.91 [0.83-0.96]** | **0.81 [0.71-0.91]** | **-0.08 [-0.14- -0.02]** | **[-0.12; -0.04]** |
| Postural Tremor | 27 | 0.90 [0.71-1] | 0.56 [0.34-0.92] | -0.17 [-0.33-0] | [-0.27; -0.02] |
| Intention Tremor | 28 | 0.95 [0.86-0.99] | 0.86 [0.76-0.95] | -0.06 [-0.15- -0.01] | [-0.11; -0.03] |
| **PD** |  |  |  |  |  |
| **Total Tremor** | **29** | **0.81 [0.65-0.93]** | **0.64 [0.31-0.83]** | **-0.10 [-0.22- -0.02]** | **[-0.20; -0.04]** |
| Rest Tremor | 29 | 0.91 [0.60-0.99] | 0.68 [0.22-0.93] | -0.16 [-0.34-0] | [-0.26; -0.04] |
| Action Tremor | 26 | 0.87 [0.61-0.95] | 0.74 [0.31-0.82] | -0.13 [-0.16-0] | [-0.15; -0.01] |
| Akinetic-rigid symptoms | 29 | 0.75 [0.60-1] | 0.50 [0.24-0.67] | -0.29 [-0.45-0] | [-0.40; 0.00] |
| **Combined** |  |  |  |  |  |
| **Total Tremor** | **57** | **0.85 [0.76-0.94]** | **0.77 [0.52-0.89]** | **-0.08 [-0.22- -0.02]** | **[-0.13; -0.05]** |
| Rest Tremor | 40 | 0.95 [0.69-1] | 0.74 [0.31-0.93] | -0.12 [-0.26- -0.03] | [-0.23; -0.07] |
| Postural Tremor | 52 | 0.88 [0.62-1] | 0.66 [0.28-0.82] | -0.15 [-0.27-0] | [-0.21; -0.05] |
| Kinetic Tremor | 44 | 0.92 [0.85-0.99] | 0.84 [0.68-0.93] | -0.06 [-0.15- -0.01] | [-0.10; -0.03] |

### Secondary Outcomes

For ET, non-inferiority of imaging-guided contact selection was shown for intention tremor but not postural tremor at the fixed amplitude (Figure 3C, Table 1). For PD, imaging-guided contact selection remained within the non-inferiority-margin for action tremor, but not for rest tremor (Figure 4C, Table 1). For akinetic-rigid symptoms, imaging guided-contact selection was inferior to clinical contact selection. Stimulation with the maximal tolerated amplitude resulted in non-inferior effects on tremor control across diseases and tremor subtypes (Supplementary Table S3). The Wilcoxon signed-rank test indicated that the side effect threshold of the best clinical contact (median ET: 3.0 mA, IQR 1.5 to 4 mA; PD: 4.5 mA, IQR 4 to 5.5 mA) was higher than that of the best imaging-guided contact (median ET: 2.5 mA, IQR 2 to 3.5 mA; PD: 4 mA, IQR 3.5 to 5.13 mA), (ET: p = 0.002, z = 3.1; PD: p = 0.012, z = 2.5, Supplementary Table S4). Clinical testing took an average of 06:11 minutes per contact in ET and 08:00 minutes in PD, resulting in an overall duration of the monopolar assessment of 80 (ET) and 95 minutes (PD) per hemisphere. Imaging-guided contact selection took approximately 30 minutes of active work by the investigator per patient for data transfer and quality control after establishing an automated imaging analysis pipeline.

A pooled analysis of both cohorts resulted in a median improvement of the accelerometric tremor total score of 85% (IQR 76% to 94%) by the best clinical contact and 77% (IQR 52% to 89%) by the best imaging-guided contact. The median difference between both contact selection strategies was-8% (IQR-22% to-2%) with a 95% CI of-13% to-5% (Table 1, Figure 5).

**Figure 5:**
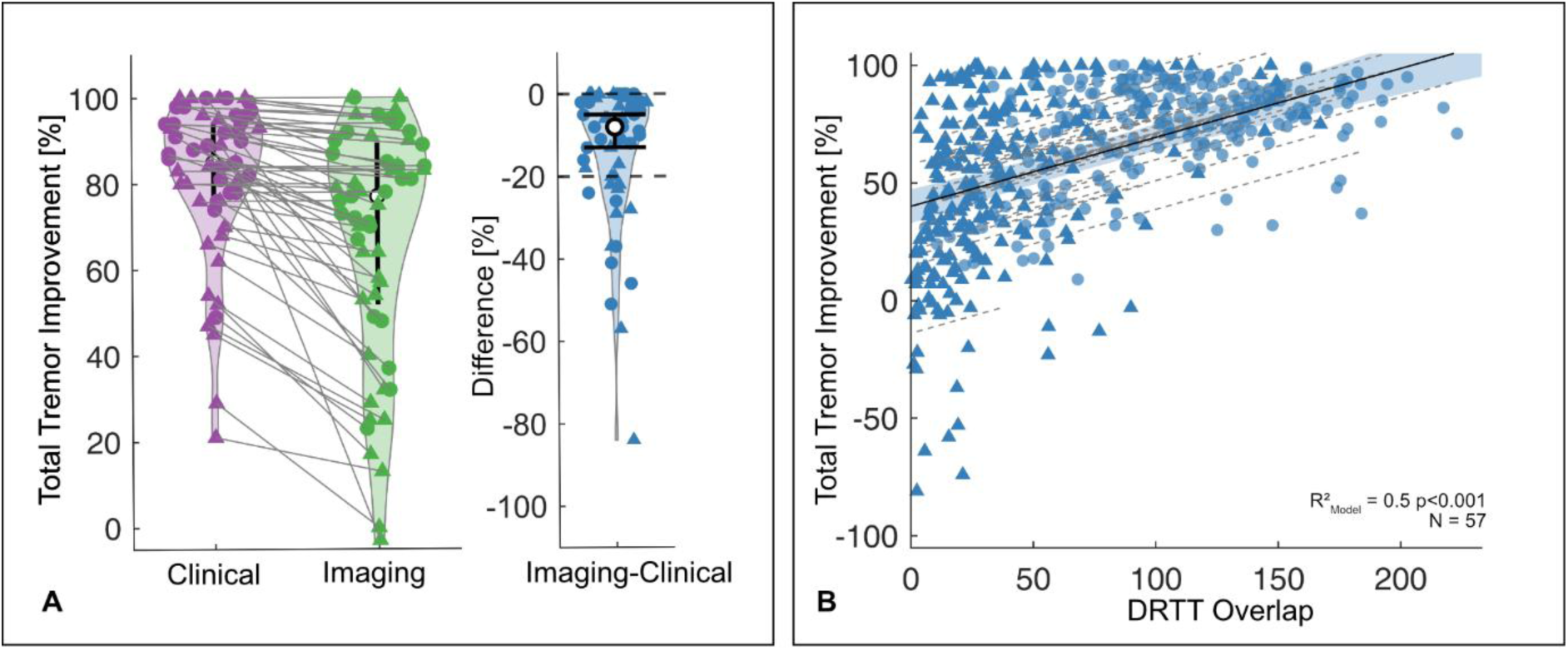
Pooled Analysis of Clinical vs. Imaging-guided Contact Selection. (A) Tremor control by stimulation via the imaging-guided selected contact was non-inferior to the best clinical contact in a pooled analysis of Essential Tremor and Parkinson’s Disease patients. (B) A moderate association between DRTT overlap and change in total tremor was observed in the combined analysis. Abbreviations: DRTT = dentato-rubro-thalamic tract

For ET, the linear mixed effects model revealed a strong association between total tremor improvement and DRTT overlap (R² model = 0.66, p<0.001, Figure 3B2, Supplementary Table S5). For PD, the association between total tremor control and overlap with the DRTT was weaker (R² model = 0.35, p<0.001, Figure 4B2, Supplementary Table S5). Similar associations were observed for all tremor subscores (Figure 3C and 4C). Of note, in PD, the effects were considerably weaker for rest tremor than for action tremor. When combining both cohorts, there was a moderate association between tremor control and DRTT overlap (R² model = 0.5, p<0.001, Figure 5, Supplementary Table S5).

## Discussion

This prospective double-blinded study provides class-II-evidence that imaging-guided contact selection is non-inferior to monopolar clinical testing for tremor suppression in DBS for ET and PD in the acute setting. This study introduces a highly standardized, feasible, and time-efficient approach to guide contact selection for tremor control by individual tractography, namely stimulation overlap with the DRTT, with the strength of an objective tremor assessment of disease-specific tremor subtypes and a study design unbiased by the potential influence of alterations of the stimulation amplitude. Our findings thereby translate previous evidence of the DRTT as a common target structure for imaging-guided contact selection across diseases into clinical practice.

### The DRTT as common target structure for imaging-guided contact selection across tremor diseases

For ET, based on normative imaging data, Dembek et al. demonstrated that the superior efficiency of PSA contacts compared to VIM contacts was related to their proximity to the DRTT [10]. Subsequent work by our group confirmed these findings utilizing individual probabilistic tractography [13]. Further studies using patient-specific and normative tracking of the DRTT have drawn similar conclusions, thereby motivating the integration of these findings into clinical practice [35–40]. Although the preliminary evidence was not as extensive as for ET, previous studies also established an association between Parkinsonian tremor control and stimulation of the DRTT [14–17]. Here, we were able to successfully apply these findings to define an imaging-guided postoperative DBS contact selection strategy for tremor control in PD, complementing previously proposed approaches for imaging-guided programming that had so far only focused on overall symptom control or akinetic-rigid symptoms [3–5,41]. In the pooled analysis of total tremor control in ET and PD, the imaging-guided approach was non-inferior to clinical testing, and there was a moderate association of the DRTT overlap in the linear mixed effect model, again supporting the hypothesis of the DRTT being a common neuromodulation key structure connecting different targets across tremor diseases [7,12].

While imaging-guided contact selection was non-inferior to the clinical approach for total tremor control in both ET and PD, we observed differences regarding tremor subtypes. In ET, non-inferiority was only established for intention but not for postural tremor. In contrast, linear effect models resulted in similar strengths of the association between the overlap with the DRTT and the achieved tremor control for both tremor subtypes (Figure 3). Therefore, the inferiority in postural tremor control might be caused by the few outliers illustrated in Figure 3C2, indicating the potential need for larger cohorts for future trials focusing on tremor subtypes. Interestingly, Deuter et al. hypothesized that only crossing fibers of the DRTT are associated with the effect of DBS on intention tremor. In contrast, postural tremor improvement was associated with both crossing and non-crossing fibers of the DRTT [42], illustrating the need for further neuroimaging analyses. In PD, non-inferiority was only observed for total tremor control and action tremor but not rest tremor. This finding was consistent with a weaker association between rest tremor and overlap with the DRTT than action tremor in the linear mixed effect models (Figure 4). The main contributing factor is most likely the fluctuation of rest tremor during testing. However, as hypothesized in prior studies, another reason might be that networks involved in the pathophysiology of rest and action tremor differ [43,44].

As expected, we found that contact selection based on the overlap with the DRTT was ineffective for improving akinetic-rigid symptoms, supporting the hypothesis of the involvement of distinct symptom-specific networks. In line with this finding, Rajamani et al. demonstrated that connections between the primary motor cortex and the most posterior region of the motor STN, as well as the decussating cerebello-thalamic tract, were associated with tremor control while fiber tracts associated with improvement of bradykinesia, axial symptoms, and rigidity were connected to the Supplementary Motor Area (SMA), pre-SMA, and premotor regions, entering the STN more anteriorly [18].

There was a marginal difference in side-effect thresholds between clinical and imaging-guided contact selection strategies. This difference is likely due to the imaging-guided approach not considering fiber tracts associated with side effects, which might need to be implemented for a comprehensive approach to imaging-guided programming in the future [31].

#### Methodological Considerations and Limitations

Although we demonstrated non-inferiority of the imaging-guided contact selection strategy within the predefined non-inferiority margin of 20%, which was established based on previous studies on the minimal clinically relevant change in motor symptoms in PD [3,34], one must consider that tremor control by the imaging-guided contact selection strategy mostly resulted in inferior but non-inferior tremor control with regard to the respective 95% CI. To interpret this finding, a key methodological consideration is that, by study design, the outcomes of both strategies were determined by the same clinical testing. Therefore, the imaging-guided contact selection strategy could only achieve outcomes that were equal or inferior to those of the best clinical contact, but not superior in the acute challenge. Long-term studies are needed to investigate the impact of this potential difference in tremor control in daily living.

Imaging-guided contact selection strategies can only be as precise as the employed imaging modalities, especially when comparing the dimensions of DBS contacts (0.5 mm to 1.5 mm) to the employed voxel size of the diffusion imaging (2 mm). The imaging protocol was chosen in line with previous studies and had to be implementable in clinical practice [12,17]. Regarding the probabilistic tracking approach, there is a vast inhomogeneity about tracking algorithms in prior studies on the impact of the DRTT on tremor control, limiting the comparability of results [38]. With MRTrixTissue3, we implemented a state-of-the-art imaging analysis software with a probabilistic fiber tracking approach, which was proven to be more sensitive and robust in detecting the DRTT than deterministic alternatives [27].

As our study investigated DBS tremor control in an acute challenge, our findings only allow for direct conclusions regarding immediate DBS effects. Considering the possibly delayed evolvement of DBS effects and the fluctuating nature of Parkinsonian rest tremor, future studies should include long-term follow-ups and continuous monitoring of tremor.

### Conclusion and Future Directions

This study provides class-II evidence for a feasible standardized imaging-guided contact selection algorithm for DBS programming for tremor control in patients with ET and PD across targets, achieving non-inferiority to burdensome and time-consuming clinical testing. Additionally, these results could have implications beyond DBS programming, e.g., potentially improving targeting of high-intensity focused ultrasound as a precision one-time treatment for tremor [45,46]. Future studies extending our pathophysiologic understanding of the modulation of different tremor subtypes by DBS and implementing novel imaging techniques such as FGATIR and fiber tracts associated with stimulation-induced side effects hold the potential to refine the proposed imaging-based programming strategy [31,47].

## Data Availability

The main outcome data supporting the findings of this study and scripts for the employed MRtrix3Tissue pipeline are available via the open science framework (https://osf.io/) at http://doi.org/10.17605/OSF.IO/34ZR2 (*data added upon publication*). Anonymized imaging data is available upon reasonable request to the corresponding author.

## CRediT authorship contribution statement

**Christina van der Linden:** Data curation, Formal analysis, Investigation, Methodology, Project administration, Software, Visualization, Writing – original draft, Writing – review & editing. **Thea Berger:** Investigation, Project administration, Writing – review & editing. **Till A. Dembek:** Conceptualization, Data curation, Formal analysis, Methodology, Software, Visualization, Writing – review & editing. **Gregor A. Brandt:** Formal analysis, Methodology, Writing – review & editing. **Joshua N. Strelow:** Software, Writing – review & editing. **Juan Carlos Baldermann:** Investigation, Writing – review & editing. **Hannah Jergas:** Investigation, Writing – review & editing. **Gereon R. Fink:** Resources, Supervision, Writing – review & editing. **Veerle Visser-Vandewalle:** Resources, Writing – review & editing. **Michael T. Barbe:** Conceptualization, Methodology, Resources, Supervision, Writing – review & editing. **Jan Niklas Petry-Schmelzer:** Conceptualization, Formal analysis, Funding acquisition, Investigation, Methodology, Supervision, Visualization, Writing – original draft, Writing – review & editing.

## Funding sources

This work was supported by the German Research Foundation (Deutsche Forschungsgemeinschaft, DFG, Project Number 502436811). Till A. Dembek and Jan Niklas Petry-Schmelzer were supported by the Cologne Clinician Scientist Program (CCSP)/ Faculty of Medicine/ University of Cologne, funded by the German Research Foundation (DFG, FI 773/15-1). Thea Berger was supported by the Koeln Fortune Program/Faculty of Medicine, University of Cologne. Juan Carlos Baldermann is funded by Else Kroener-Fresenius-Stiftung (2022_EKES.23) and receives funding from the German Research Foundation (Project ID 431549029-C07). Hannah Jergas received funding by Koeln Fortune, FMMED and EIT Health.

## Declaration of competing interests

Till A. Dembek, Gregor A. Brandt, Joshua N. Strelow, Hannah Jergas, Veerle Visser-Vandewalle and Michael T. Barbe have business relations with or received funding from at least one of Medtronic, Abbott, and Boston Scientific, which produce DBS devices, but none was related to the current work. Gereon R. Fink received royalties from the publication of the books *Funktionelle MRT in Psychiatrie und Neurologie*, *Neurologische Differentialdiagnose*, *SOP Neurologie*, and *Therapiehandbuch Neurologie* and from the publication of the neuropsychological tests *KAS, NP-KiSS*, and *KöpSS*, honoraria for speaking engagements from the Deutsche Gesellschaft für Neurologie (DGN) and Forum für medizinische Fortbildung FomF GmbH. Michael T. Barbe was supported by Felgenhauer-Stiftung, Forschungspool Klinische Studien, received speakers honoraria from Bial, Medtronic, Boston Scientific, Abbott, FomF, GE medical, UCB, Bund Deutscher Neurologen, Esteve, Apothekerverband Köln e.V. as well as advisory honoraria for the IQWIG, Medtronic, Esteve and Abbvie.

## Supplementary Material

### 1 Supplementary Methods

#### 1.1 Accelerometry

For recording the accelerometric data, we attached a BrainProducts (BrainProducts GmbH, Gilching, Germany) triaxial accelerometer to the proximal dorsum of the index finger (Sampling rate 2500 Hz; ±2 g acceleration sensing range; <0.001 g resolution) and a triaxial wristwatch accelerometer (GENEActiv© Original, ActivinsightsTM, Kimbolton, UK; sampling rate 100 Hz; ±8 g acceleration sensing range; 0.004 g resolution) to the wrist. For rest and postural tremor data, pre-processing was performed as described in van der Linden et al. [1]. In this prior work, we showed that of four evaluated metrics for tremor quantification, Mean Acceleration, calculated by summating all absolute acceleration values within the filtered acceleration data and divided by the number of samples, measured at the finger, was the metric with the highest interrater-reliability (weighted kappa and percentage concordance) with clinical rating. We calculated an accelerometric tremor score presuming a non-linear relationship between clinical ratings and accelerometry, illustrated by the equation *TR = a ∗ T^c^ + b*, wherein *T* is the tremor severity assessed by Mean Acceleration, *TR* the tremor rating score, *a* the slope, *b* the intercept, and *c* the exponent of a power relationship between *T* and *TR*. Optimal model parameters for *a*, *b* and *c,* determined by a non-linear regression analysis in a training data-set with validation in a holdout dataset before [1], were applied for tremor score calculations for each rest and postural tremor trial. For the accelerometric assessment of intention/kinetic tremor, we used the same preprocessing steps but divided each 20s trial into subsections based on the timestamps recorded when patients pressed the button, representing the endpoint of the intention movement. To focus on the intention tremor component, only the last 25% of the duration between two recorded timestamps were considered, and the average Mean Acceleration of all subsections was calculated. Applying the previously described methodology^1^, we determined optimal model parameters for the non-linear model in the training data set and validated them in the remaining data (Finger [Wrist]: weighted kappa = 0.5 [0.4], percentage concordance of 67.4% [63.1%], Table S1). We employed Mean Acceleration measured at the finger for all but three hemispheres (all PD), where we employed wrist data due to missing values.

#### 1.2 Diffusion Imaging Analysis

All patients underwent routine MR-imaging for surgical planning. MRI data were acquired on a 3-Tesla Philips Ingenia®Scanner (Philips, Amsterdam, The Netherlands). For the present study, we employed the T1-sequence (TR: 9.8 ms; TE: 4.9 ms; acquisition time: 6.13 min; voxel-size: 0.49 × 0.49 × 1.00mm³) and the diffusion imaging sequence (TR: 8213 ms; TE: 103 ms; 40 gradient directions; b-value: 1000 s/mm²; acquisition time: 9:53 min; voxel-size; 2.0 × 2.0 × 2.0 mm). Diffusion data processing was performed using the MRtrix3Tissue package (https://3Tissue.github.io), a fork of MRtrix3 [2]. For preprocessing, diffusion data was (i) denoised^21^ and unringed to remove Gibb’s ringing artifacts [3], (ii) corrected for susceptibility-induced distortions and head motion, using the topup and eddy tool from the FMRIB software library (FMRIB, Oxford, UK), and (iii) bias field corrected as implemented in Advanced Normalization Tools (ANTS) [4]. Brain masks were created using the SynthStrip Tool [5]. Then, single-shell 3-Tissue constrained spherical devolution was performed to obtain white-matter-like fiber orientation distributions and grey-matter-like and cerebrospinal fluid-like compartments in all voxels [6,7]. Hereby, we effectively addressed the challenge of tracking crossing fibers of the decussating DRTT [8]. Probabilistic fiber tracking of the DRTT was conducted using the contralateral dentate nucleus as the seed region, while the contralateral superior cerebellar peduncle, the ipsilateral red nucleus, and the ipsilateral precentral gyrus served as waypoints (Manuscript Figure 1B). Regions of interest were delineated in the MNI ICBM 2009b asym. template to ensure a standardized tracking result without inter-rater variability [9]. Afterwards, they were transformed to the preoperative T1 space using ANTs, and further co-registered to the averaged b0 using SPM (http://www.fil.ion.ucl.ac.uk/spm/software/spm12/), as described previously [10,11]. The iFOD2-algorithm [12] was conducted with 200, 500, and 1000 selected fibers respectively, unlimited seeds, a default step size of 0.5 x voxel size, a minimum length of any tract of 10 mm, a maximum length of any tract of 300 mm and unidirectional tracking. Following visual inspection about plausibility and inter-individual comparability, only tracts with 500 selected fibers were included for further analysis. Finally, the resulting tracts were co-registered to the preoperative T1, based on the previously derived co-registration matrix, so that the overlap of the individual DRTT with the local stimulation spread could be calculated in the individual T1 space.

#### 1.3 Calculation of Contact-Wise DRTT Overlap

DBS leads were reconstructed from postoperative CT scans and transformed into T1 space utilizing a well-established workflow in the LEAD-DBS toolbox (version 3, www.lead-dbs.org) [13–15]. Postoperative CT images (IQon Spectral CT, iCT 256, Brilliance 256, Philips Healthcare, Best, The Netherlands) were linearly co-registered to preoperative magnetic resonance imaging (3-Tesla Philips Ingenia® Scanner, Philips, Amsterdam, The Netherlands) using advanced normalization tools (ANTs, http://stnava.github.io/ANTs/) [16]. A brain shift correction step was applied as implemented in Lead-DBS. Then, images were non-linearly normalized into standard space (ICBM 2009b NLIN asym.) using ANTs and the “Effective: low variance, Default” setting. The results of each pre-processing step were visually inspected regarding accuracy and refined if necessary. DBS leads were automatically reconstructed with the PaCER algorithm [17], and manually refined. The orientation of the directional leads was determined using the DiODE algorithm [18]. The local stimulation spread at the fixed amplitude was calculated in T1 space for each contact. To do so, precomputed electric fields were employed for each stimulation setting to estimate the spread of the electric field for homogenous tissue with a conductivity of σ = 0.2 S/m (FastField) [19–21]. Instead of applying a fixed electric field threshold resulting in a binarized stimulation volume, a recently established probabilistic stimulation model based on a sigmoidal activation function and established electric field activation thresholds for different fiber diameters was used, resulting in a voxel-wise activation probability value reaching from 0 (= no activation) to asymptotic 1 [22]. Ultimately, the overlap of each stimulation spread with the respective DRTT was calculated as the sum of the fiber-wise maximal activation probability of each fiber, stimulated with an activation probability of at least 0.05. Contacts were ranked lead-wise by overlap, declaring the contact with the highest overlap as the best imaging contact. Investigators performing the tracking and overlap analysis (JNPS) were blinded to the clinical testing results and vice versa.

## Data Availability

The main outcome data supporting the findings of this study and scripts for the employed MRtrix3Tissue pipeline will be made available via the open science framework (https://osf.io/) at http://doi.org/10.17605/OSF.IO/34ZR2 upon publication. Anonymized imaging data is available upon reasonable request to the corresponding author.

## 2 Supplementary Tables

**Table S1:** Accelerometry Model Parameters for Intention/Kinetic Tremor. Model parameters (coefficients a, b, and c) determined in the training dataset, weighted Kappa and Concordance between clinical ratings and the resulting accelerometric tremor score when applying the model parameters to the validation data set. Abbreviations: *a* = slope, *b* = intercept, *c* = exponent of a power relationship between *T* and *TR*, RMSE = root mean squared error, T = tremor severity assessed by Mean Acceleration, TR = tremor rating score

| Location | TR = a * T<sup>c</sup> + b |  |  | RMSE | Weighted Kappa | Concordance |
| --- | --- | --- | --- | --- | --- | --- |
|  | a | b | c |  |  |  |
| Finger | 4.47 | -0.39 | 0.94 | 0.628 | 0.50 | 0.674 |
| Wrist | 8.91 | -0.29 | 1.11 | 0.917 | 0.40 | 0.631 |

**Table S2:**
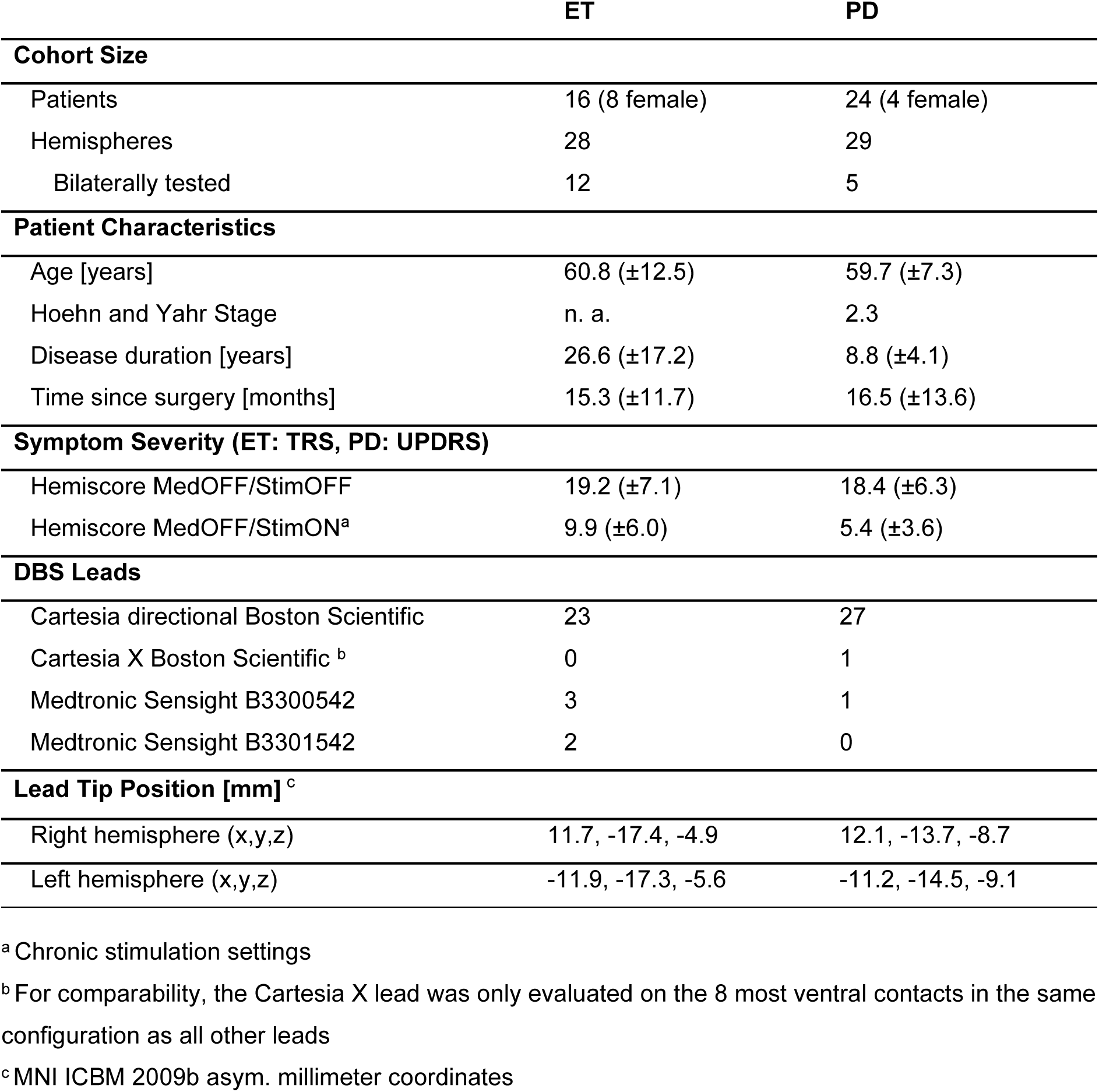
Patient Characteristics. Abbreviations: DBS = deep brain stimulation, ET = Essential Tremor, MedOFF = after 12 hours withdrawal of disease-specific medication, PD = Parkinson’s Disease, StimOFF = Stimulation OFF, StimON = Stimulation ON, TRS = Fahn Tolosa Marin Tremor Rating Scale, UPDRS = Unified Parkinson’s Disease Rating Scale

**Table S3:** Tremor Improvement at Maximal Amplitude. Median percentage improvement of the accelerometric tremor score achieved by the best clinical contact and the best imaging-guided contact with the maximal amplitude (0.5 mA below the occurrence of side effects) and median of the differences between the improvement on the best clinical vs. the best imaging contact. Abbreviations: CI = Confidence Interval, ET = Essential Tremor, IQR = interquartile range, PD = Parkinson’s Disease

| Outcome parameter | N | Median Improvement Clinical [IQR] | Median Improvement Imaging [IQR] | Median Differences Clinical – Imaging [IQR] | 95 % CI |
| --- | --- | --- | --- | --- | --- |
| **ET** |  |  |  |  |  |
| **Total Tremor** | **28** | **0.92 [0.83-0.96]** | **0.88 [0.74-0.94]** | **-0.02 [-0.11-0]** | **[-0.07; 0]** |
| Postural Tremor | 27 | 0.92 [0.73-1] | 0.84 [0.52-0.99] | 0 [-0.11-0] | [-0.07; 0] |
| Intention Tremor | 28 | 0.96 [0.89-1] | 0.92 [0.85-0.97] | -0.03 [-0.05-0] | [-0.05; 0] |
| **PD** |  |  |  |  |  |
| **Total Tremor** | **29** | **0.83 [0.64-0.92]** | **0.76 [0.59-0.88]** | **-0.03 [-0.13-0]** | **[-0.09; 0]** |
| Rest Tremor | 29 | 0.86 [0.68-0.99] | 0.77 [0.56-0.9] | -0.01 [-0.13-0] | [-0.11; 0] |
| Action Tremor | 26 | 0.77 [0.55-0.9] | 0.78 [0.45-0.9] | 0 [-0.05-0.02] | [-0.04; 0.01] |
| Akinetic-rigid Symptoms | 29 | 0.86 [0.75-1] | 0.67 [0.5-1.0] | 0 [0.33-0] | [-0.17; 0] |
| **Combined** |  |  |  |  |  |
| **Total Tremor** | **57** | **0.88 [0.77-0.95]** | **0.8 [0.66-0.92]** | **-0.02 [-0.11-0]** | **[-0.05; 0]** |
| Rest Tremor | 40 | 0.91 [0.72-1] | 0.78 [0.55-0.92] | -0.07 [-0.17-0] | [-0.11; 0] |
| Postural Tremor | 52 | 0.84 [0.62-0.95] | 0.78 [0.48-0.93] | 0 [-0.11-0] | [-0.04; 0] |
| Kinetic Tremor | 44 | 0.94 [0.81-0.99] | 0.89 [0.76-0.97] | -0.01 [-0.05-0] | [-0.03; 0] |

**Table S4:** Side Effect Thresholds of the Best Clinical vs. Imaging Contact (Total Tremor). Median side effect thresholds of the most effective clinical contact and the imaging-guided contact. Abbreviations: ET = Essential Tremor, PD = Parkinson’s Disease, p-and z-Values as indicated by a Wilcoxon signed-rank test.

|  | N | Median threshold Clinical [IQR] in mA | Median threshold Imaging [IQR] in mA | p | z | Median difference |
| --- | --- | --- | --- | --- | --- | --- |
| ET | 28 | 3.0 [1.5-4] | 2.5 [2-3.25] | 0.0021 | 3.07 | -0.5 [-1-0] |
| PD | 29 | 4.5 [4-5.5] | 4.0 [3.5-5.13] | 0.0122 | 2.5 | 0 [-1-0] |

**Table S5:** Association between Tremor Improvement and Overlap with the DRTT. Results of the Linear Mixed Effect Model with “individual DRTT overlap” as the fixed effect and lead as the random effect for Essential Tremor, Parkinson’s Disease and the combined dataset. Abbreviations: ET = Essential Tremor, PD = Parkinson’s Disease

|  | N | R<sup>2</sup> model | p |
| --- | --- | --- | --- |
| **ET** |  |  |  |
| Total Tremor | 28 | 0.661 | <0.001 |
| Postural Tremor | 27 | 0.547 | <0.001 |
| Intention Tremor | 28 | 0.686 | <0.001 |
| **PD** |  |  |  |
| Total Tremor | 29 | 0.351 | <0.001 |
| Rest Tremor | 29 | 0.277 | <0.001 |
| Action Tremor | 26 | 0.699 | <0.001 |
| Akinetic-rigid Symptoms | 29 | 0.247 | 0.008 |
| **Combined** |  |  |  |
| Total Tremor | 57 | 0.502 | <0.001 |
| Rest Tremor | 40 | 0.316 | <0.001 |
| Postural Tremor | 52 | 0.695 | <0.001 |
| Kinetic Tremor | 44 | 0.752 | <0.001 |

## 3 Supplementary Figures

**Supplementary Figure S1:**
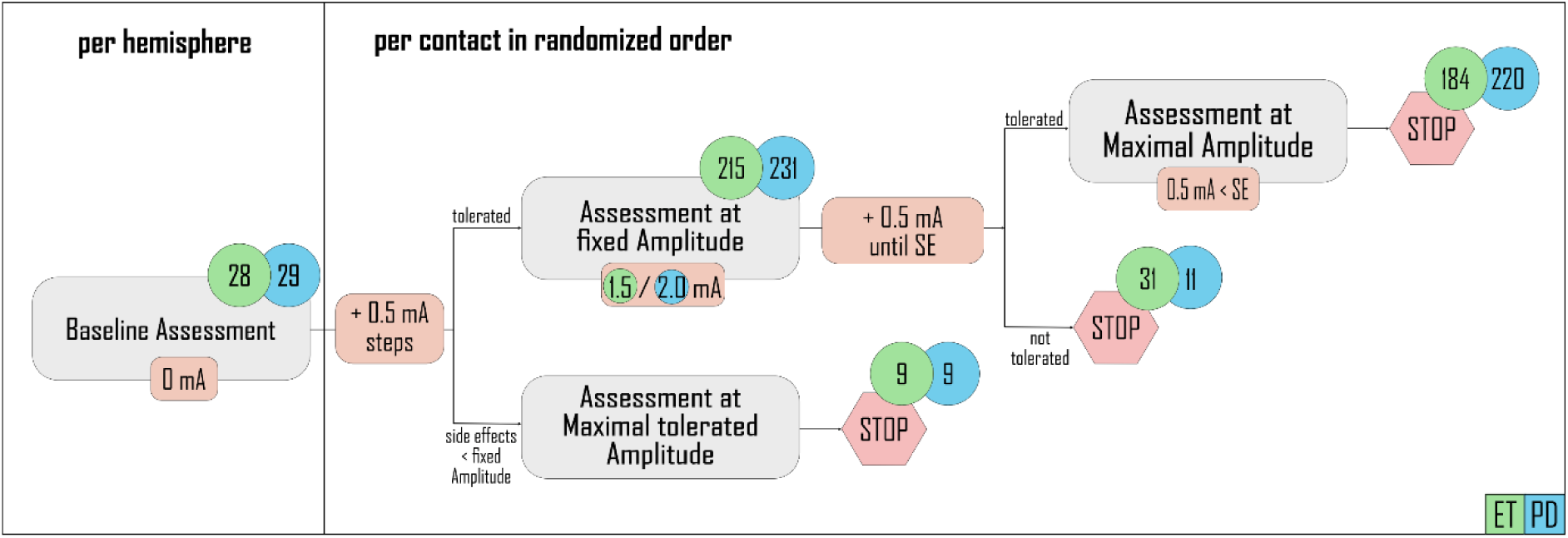
Course of Clinical Testing. In patients with Essential Tremor [green], the fixed amplitude could be reached without side effects on 215 contacts. Side effects occurred 0.5 mA above the fixed amplitude on 31 contacts, allowing only one measurement at the fixed amplitude of 1.5 mA. This resulted in 184 contacts with both measurements [A] and [B] possible, 31 contacts only with measurement [A] (1.5 mA), and 9 contacts where the fixed amplitude could not be reached, so that only measurement [B] was conducted 0.5 mA below the side effect threshold. In patients with Parkinson’s Disease [blue], the fixed amplitude of 2 mA was reached without side effects on 231 contacts, with 220 contacts where side effects occurred > 0.5 mA above the fixed amplitude of 2.0 mA, making both measurements possible, 11 contacts where side effects occurred at 2.5 mA so that only measurement [A] was taken, and 9 contacts with side effects occurring < 2 mA, so that only measurement [B] was possible. For the primary outcome (improvement of the total accelerometric tremor score), the best clinical contact was determined based on the effect at the fixed amplitude in 50 cases, by the effect at maximal amplitude in four cases, and by the highest side-effect threshold in two cases. For one lead, there were two best clinical contacts, with 100% tremor reduction at both fixed and maximal amplitude and equal side effect thresholds. Abbreviations: ET = Essential Tremor, PD = Parkinson’s Disease, SE = side effects

## Notes

### Clinical Trial

DRKS00026596

### Author Declarations

The ethics committee of the University of Cologne gave ethical approval for this study (21-1441), conducted under the Declaration of Helsinki, and registered with the German Clinical Trials Registry before patient recruitment (DRKS00026596). All patients gave written informed consent before participation.

## References

[1] Barbe MT, Reker P, Hamacher S, Franklin J, Kraus D, Dembek TA, et al. DBS of the PSA and the VIM in essential tremor: A randomized, double-blind, crossover trial. Neurology 2018;91:e543–50. 10.1212/WNL.0000000000005956.

[2] Fasano A, Daniele A, Albanese A. Treatment of motor and non-motor features of Parkinson’s disease with deep brain stimulation. Lancet Neurol 2012;11:429–42. 10.1016/S1474-4422(12)70049-2.

[3] Waldthaler J, Bopp M, Kühn N, Bacara B, Keuler M, Gjorgjevski M, et al. Imaging-based programming of subthalamic nucleus deep brain stimulation in Parkinson’s disease. Brain Stimulat 2021;14:1109–17. 10.1016/j.brs.2021.07.064.

[4] Brandt GA, Stopic V, Van Der Linden C, Strelow JN, Petry-Schmelzer JN, Baldermann JC, et al. A Retrospective Comparison of Multiple Approaches to Anatomically Informed Contact Selection in Subthalamic Deep Brain Stimulation for Parkinson’s Disease. J Park Dis 2024:1–13. 10.3233/JPD-230200.

[5] Lange F, Steigerwald F, Malzacher T, Brandt GA, Odorfer TM, Roothans J, et al. Reduced Programming Time and Strong Symptom Control Even in Chronic Course Through Imaging-Based DBS Programming. Front Neurol 2021;12:785529.

[6] Buijink AWG, van Rootselaar A-F, Helmich RC. Connecting tremors - a circuits perspective. Curr Opin Neurol 2022;35:518–24. 10.1097/WCO.0000000000001071.

[7] Fox MD, Deuschl G. Converging on a Neuromodulation Target for Tremor. Ann Neurol 2022;91:581–4. 10.1002/ana.26361.

[8] Meola A, Comert A, Yeh F-C, Sivakanthan S, Fernandez-Miranda JC. The nondecussating pathway of the dentatorubrothalamic tract in humans: human connectome-based tractographic study and microdissection validation. J Neurosurg 2016;124:1406–12. 10.3171/2015.4.JNS142741.

[9] Kvernmo N, Konglund AE, Reich MM, Roothans J, Pripp AH, Dietrichs E, et al. Deep Brain Stimulation for Arm Tremor: A Randomized Trial Comparing Two Targets. Ann Neurol 2022;91:585–601. 10.1002/ana.26317.

[10] Dembek TA, Petry-Schmelzer JN, Reker P, Wirths J, Hamacher S, Steffen J, et al. PSA and VIM DBS efficiency in essential tremor depends on distance to the dentatorubrothalamic tract. NeuroImage Clin 2020;26:102235. 10.1016/j.nicl.2020.102235.

[11] Fenoy AJ, Schiess MC. Deep Brain Stimulation of the Dentato-Rubro-Thalamic Tract: Outcomes of Direct Targeting for Tremor. Neuromodulation Technol Neural Interface 2017;20:429–36. 10.1111/ner.12585.

[12] Coenen VA, Allert N, Paus S, Kronenbürger M, Urbach H, Mädler B. Modulation of the Cerebello-Thalamo-Cortical Network in Thalamic Deep Brain Stimulation for Tremor: A Diffusion Tensor Imaging Study. Neurosurgery 2014;75:657. 10.1227/NEU.0000000000000540.

[13] Petry-Schmelzer J, Dembek T, Steffen J, Jergas H, Dafsari H, Fink G, et al. Selecting the Most Effective DBS Contact in Essential Tremor Patients Based on Individual Tractography. Brain Sci 2020;10:1015. 10.3390/brainsci10121015.

[14] Abdulbaki A, Kaufmann J, Galazky I, Buentjen L, Voges J. Neuromodulation of the subthalamic nucleus in Parkinson’s disease: the effect of fiber tract stimulation on tremor control. Acta Neurochir (Wien) 2021;163:185–95. 10.1007/s00701-020-04495-3.

[15] Prent N, Potters WV, Boon LI, Caan MWA, De Bie RMA, Van Den Munckhof P, et al. Distance to white matter tracts is associated with deep brain stimulation motor outcome in Parkinson’s disease. J Neurosurg 2020;133:433–42. 10.3171/2019.5.JNS1952.

[16] Helmich RC, Hallett M, Deuschl G, Toni I, Bloem BR. Cerebral causes and consequences of parkinsonian resting tremor: a tale of two circuits? Brain 2012;135:3206–26. 10.1093/brain/aws023.

[17] Coenen VA, Sajonz B, Prokop T, Reisert M, Piroth T, Urbach H, et al. The dentato-rubro-thalamic tract as the potential common deep brain stimulation target for tremor of various origin: an observational case series. Acta Neurochir (Wien) 2020;162:1053–66. 10.1007/s00701-020-04248-2.

[18] Rajamani N, Friedrich H, Butenko K, Dembek T, Lange F, Navrátil P, et al. Deep brain stimulation of symptom-specific networks in Parkinson’s disease. Nat Commun 2024;15:4662. 10.1038/s41467-024-48731-1.

[19] Deuschl G, Bain P, Brin M, Committee AHS. Consensus Statement of the Movement Disorder Society on Tremor. Mov Disord 1998;13:2–23. 10.1002/mds.870131303.

[20] Postuma RB, Berg D, Stern M, Poewe W, Olanow CW, Oertel W, et al. MDS clinical diagnostic criteria for Parkinson’s disease: MDS-PD Clinical Diagnostic Criteria. Mov Disord 2015;30:1591–601. 10.1002/mds.26424.

[21] Fahn S, Tolosa E, Marin C. Clinical Rating Scale for Tremor. Park. Dis. Mov. Disord., Baltimore: Williams and Wilkins; 1993, p. 271–80.

[22] Goetz CG, Tilley BC, Shaftman SR, Stebbins GT, Fahn S, Martinez-Martin P, et al. Movement Disorder Society-sponsored revision of the Unified Parkinson’s Disease Rating Scale (MDS-UPDRS): Scale presentation and clinimetric testing results. Mov Disord 2008;23:2129–70. 10.1002/mds.22340.

[23] Thompson JA, Hirt L, David-Gerecht P, Fasano A, Kramer DR, Ojemann SG, et al. Comparison of Monopolar Review to Fixed Parameter Fractionation in Deep Brain Stimulation. Mov Disord Clin Pract 2023;10:987–91. 10.1002/mdc3.13750.

[24] Brainard DH. The Psychophysics Toolbox. Spat Vis 1997;10:433–6.

[25] Pelli DG. The VideoToolbox software for visual psychophysics: transforming numbers into movies. Spat Vis 1997;10:437–42. 10.1163/156856897X00366.

[26] Tournier J-D, Smith R, Raffelt D, Tabbara R, Dhollander T, Pietsch M, et al. *MRtrix3*: A fast, flexible and open software framework for medical image processing and visualisation. NeuroImage 2019;202:116137. 10.1016/j.neuroimage.2019.116137.

[27] Schlaier JR, Beer AL, Faltermeier R, Fellner C, Steib K, Lange M, et al. Probabilistic vs. deterministic fiber tracking and the influence of different seed regions to delineate cerebellar-thalamic fibers in deep brain stimulation. Eur J Neurosci 2017;45:1623–33. 10.1111/ejn.13575.

[28] Neudorfer C, Butenko K, Oxenford S, Rajamani N, Achtzehn J, Goede L, et al. Lead-DBS v3.0: Mapping deep brain stimulation effects to local anatomy and global networks. NeuroImage 2023;268:119862. 10.1016/j.neuroimage.2023.119862.

[29] Baniasadi M, Proverbio D, Gonçalves J, Hertel F, Husch A. FastField: An open-source toolbox for efficient approximation of deep brain stimulation electric fields. NeuroImage 2020;223:117330. 10.1016/j.neuroimage.2020.117330.

[30] Astrom M, Diczfalusy E, Martens H, Wardell K. Relationship between neural activation and electric field distribution during deep brain stimulation. IEEE Trans Biomed Eng 2015;62:664–72. 10.1109/TBME.2014.2363494.

[31] Jergas H, Petry-Schmelzer JN, Hannemann JH, Thies T, Strelow JN, Rubi-Fessen I, et al. One side effect: two networks? Lateral and posteromedial stimulation spreads induce dysarthria in subthalamic deep brain stimulation for Parkinson’s disease. J Neurol Neurosurg Psychiatry 2024:jnnp-2024-333434. 10.1136/jnnp-2024-333434.

[32] Van Der Linden C, Berger T, Brandt GA, Strelow JN, Jergas H, Baldermann JC, et al. Accelerometric Classification of Resting and Postural Tremor Amplitude. Sensors 2023;23:8621. 10.3390/s23208621.

[33] Waldthaler J, Bopp M, Kühn N, Bacara B, Keuler M, Gjorgjevski M, et al. Imaging-based programming of subthalamic nucleus deep brain stimulation in Parkinson’s disease. Brain Stimulat 2021;14:1109–17. 10.1016/j.brs.2021.07.064.

[34] Schrag A, Sampaio C, Counsell N, Poewe W. Minimal clinically important change on the unified Parkinson’s disease rating scale. Mov Disord 2006;21:1200–7. 10.1002/mds.20914.

[35] Avecillas-Chasin JM, Honey CR, Heran MKS, Krüger MT. Sweet spots of standard and directional leads in patients with refractory essential tremor: white matter pathways associated with maximal tremor improvement. J Neurosurg 2022;137:1811–20. 10.3171/2022.3.JNS212374.

[36] Tsuboi T, Wong JK, Eisinger RS, Okromelidze L, Burns MR, Ramirez-Zamora A, et al. Comparative connectivity correlates of dystonic and essential tremor deep brain stimulation. Brain 2021;144:1774–86. 10.1093/brain/awab074.

[37] Nowacki A, Barlatey S, Al-Fatly B, Dembek T, Bot M, Green AL, et al. Probabilistic mapping reveals optimal stimulation site in essential tremor. Ann Neurol 2022:ana.26324. 10.1002/ana.26324.

[38] Middlebrooks EH, Okromelidze L, Carter RE, Jain A, Lin C, Westerhold E, et al. Directed stimulation of the dentato-rubro-thalamic tract for deep brain stimulation in essential tremor: a blinded clinical trial. Neuroradiol J 2022;35:203–12. 10.1177/19714009211036689.

[39] Neudorfer C, Kultas-Ilinsky K, Ilinsky I, Paschen S, Helmers A-K, Cosgrove GR, et al. The role of the motor thalamus in deep brain stimulation for essential tremor. Neurotherapeutics 2024:e00313. 10.1016/j.neurot.2023.e00313.

[40] Coenen VA, Rijntjes M, Prokop T, Piroth T, Amtage F, Urbach H, et al. One-pass deep brain stimulation of dentato-rubro-thalamic tract and subthalamic nucleus for tremor-dominant or equivalent type Parkinson’s disease. Acta Neurochir (Wien) 2016;158:773–81. 10.1007/s00701-016-2725-4.

[41] Roediger J, Dembek TA, Wenzel G, Butenko K, Kühn AA, Horn A. StimFit—A Data-Driven Algorithm for Automated Deep Brain Stimulation Programming. Mov Disord 2022;37:574–84. 10.1002/mds.28878.

[42] Deuter D, Torka E, Kohl Z, Schmidt N-O, Schlaier J. Mediation of Tremor Control by the Decussating and Nondecussating Part of the Dentato-Rubro-Thalamic Tract in Deep Brain Stimulation in Essential Tremor: Which Part Should Be Stimulated? Neuromodulation Technol Neural Interface 2023;26:1668–79. 10.1016/j.neurom.2022.04.040.

[43] Helmich RC, Van den Berg KRE, Panyakaew P, Cho HJ, Osterholt T, McGurrin P, et al. Cerebello-Cortical Control of Tremor Rhythm and Amplitude in Parkinson’s Disease. Mov Disord 2021;36:1727–9. 10.1002/mds.28603.

[44] Ni Z, Pinto AD, Lang AE, Chen R. Involvement of the cerebellothalamocortical pathway in Parkinson disease. Ann Neurol 2010;68:816–24. 10.1002/ana.22221.

[45] Krishna V, Sammartino F, Agrawal P, Changizi BK, Bourekas E, Knopp MV, et al. Prospective Tractography-Based Targeting for Improved Safety of Focused Ultrasound Thalamotomy. Neurosurgery 2019;84:160. 10.1093/neuros/nyy020.

[46] Feltrin FS, Chopra R, Pouratian N, Elkurd M, El-Nazer R, Lanford L, et al. Focused ultrasound using a novel targeting method four-tract tractography for magnetic resonance–guided high-intensity focused ultrasound targeting. Brain Commun 2022;4:fcac273. 10.1093/braincomms/fcac273.

[47] Neudorfer C, Kroneberg D, Al-Fatly B, Goede L, Kübler D, Faust K, et al. Personalizing Deep Brain Stimulation Using Advanced Imaging Sequences. Ann Neurol 2022;91:613–28. 10.1002/ana.26326.

## Supplementary References

[1] Van Der Linden C, Berger T, Brandt GA, Strelow JN, Jergas H, Baldermann JC, et al. Accelerometric Classification of Resting and Postural Tremor Amplitude. Sensors 2023;23:8621. 10.3390/s23208621.

[2] Tournier J-D, Smith R, Raffelt D, Tabbara R, Dhollander T, Pietsch M, et al. *MRtrix3*: A fast, flexible and open software framework for medical image processing and visualisation. NeuroImage 2019;202:116137. 10.1016/j.neuroimage.2019.116137.

[3] Kellner E, Dhital B, Kiselev VG, Reisert M. Gibbs-ringing artifact removal based on local subvoxel-shifts. Magn Reson Med 2016;76:1574–81. 10.1002/mrm.26054.

[4] Tustison NJ, Avants BB, Cook PA, Yuanjie Zheng, Egan A, Yushkevich PA, et al. N4ITK: Improved N3 Bias Correction. IEEE Trans Med Imaging 2010;29:1310–20. 10.1109/TMI.2010.2046908.

[5] Hoopes A, Mora JS, Dalca AV, Fischl B, Hoffmann M. SynthStrip: skull-stripping for any brain image. NeuroImage 2022;260:119474. 10.1016/j.neuroimage.2022.119474.

[6] Dhollander T, Mito R, Raffelt D, Connelly A. Improved white matter response function estimation for 3-tissue constrained spherical deconvolution 2019.

[7] Dhollander T, Connelly A. A novel iterative approach to reap the benefits of multi-tissue CSD from just single-shell (+b=0) diffusion MRI data 2016.

[8] Tournier J-D, Calamante F, Connelly A. Robust determination of the fibre orientation distribution in diffusion MRI: Non-negativity constrained super-resolved spherical deconvolution. NeuroImage 2007;35:1459–72. 10.1016/j.neuroimage.2007.02.016.

[9] Fonov V, Evans AC, Botteron K, Almli CR, McKinstry RC, Collins DL, et al. Unbiased average age-appropriate atlases for pediatric studies. NeuroImage 2011;54:313–27. 10.1016/j.neuroimage.2010.07.033.

[10] Petry-Schmelzer J, Dembek T, Steffen J, Jergas H, Dafsari H, Fink G, et al. Selecting the Most Effective DBS Contact in Essential Tremor Patients Based on Individual Tractography. Brain Sci 2020;10:1015. 10.3390/brainsci10121015.

[11] Dembek TA, Petry-Schmelzer JN, Reker P, Wirths J, Hamacher S, Steffen J, et al. PSA and VIM DBS efficiency in essential tremor depends on distance to the dentatorubrothalamic tract. NeuroImage Clin 2020;26:102235. 10.1016/j.nicl.2020.102235.

[12] Tournier J-D, Calamante F, Connelly A. Improved probabilistic streamlines tractography by 2nd order integration over fibre orientation distributions n.d.

[13] Horn A, Kühn AA. Lead-DBS: A toolbox for deep brain stimulation electrode localizations and visualizations. NeuroImage 2015;107:127–35. 10.1016/j.neuroimage.2014.12.002.

[14] Horn A, Li N, Dembek TA, Kappel A, Boulay C, Ewert S, et al. Lead-DBS v2: Towards a comprehensive pipeline for deep brain stimulation imaging. NeuroImage 2019;184:293–316. 10.1016/j.neuroimage.2018.08.068.

[15] Neudorfer C, Butenko K, Oxenford S, Rajamani N, Achtzehn J, Goede L, et al. Lead-DBS v3.0: Mapping deep brain stimulation effects to local anatomy and global networks. NeuroImage 2023;268:119862. 10.1016/j.neuroimage.2023.119862.

[16] Avants BB, Tustison NJ, Song G, Cook PA, Klein A, Gee JC. A reproducible evaluation of ANTs similarity metric performance in brain image registration. NeuroImage 2011;54:2033–44. 10.1016/j.neuroimage.2010.09.025.

[17] Husch A, V. Petersen M, Gemmar P, Goncalves J, Hertel F. PaCER - A fully automated method for electrode trajectory and contact reconstruction in deep brain stimulation. NeuroImage Clin 2018;17:80–9. 10.1016/j.nicl.2017.10.004.

[18] Dembek TA, Hoevels M, Hellerbach A, Horn A, Petry-Schmelzer JN, Borggrefe J, et al. Directional DBS leads show large deviations from their intended implantation orientation. Parkinsonism Relat Disord 2019;67:117–21. 10.1016/j.parkreldis.2019.08.017.

[19] Horn A, Reich M, Vorwerk J, Li N, Wenzel G, Fang Q, et al. Connectivity Predicts deep brain stimulation outcome in Parkinson disease. Ann Neurol 2017;82:67–78. 10.1002/ana.24974.

[20] Baniasadi M, Proverbio D, Gonçalves J, Hertel F, Husch A. FastField: An open-source toolbox for efficient approximation of deep brain stimulation electric fields. NeuroImage 2020;223:117330. 10.1016/j.neuroimage.2020.117330.

[21] Astrom M, Diczfalusy E, Martens H, Wardell K. Relationship between neural activation and electric field distribution during deep brain stimulation. IEEE Trans Biomed Eng 2015;62:664–72. 10.1109/TBME.2014.2363494.

[22] Jergas H, Petry-Schmelzer JN, Hannemann JH, Thies T, Strelow JN, Rubi-Fessen I, et al. One side effect: two networks? Lateral and posteromedial stimulation spreads induce dysarthria in subthalamic deep brain stimulation for Parkinson’s disease. J Neurol Neurosurg Psychiatry 2024:jnnp-2024-333434. 10.1136/jnnp-2024-333434.

